# Can Multimodal Large Language Models Visually Interpret Auditory Brainstem Responses?

**DOI:** 10.64898/2026.04.15.26350944

**Authors:** W. Wiktor Jedrzejczak, Krzysztof Kochanek, Henryk Skarżyński

## Abstract

**Introduction:** Auditory brainstem response (ABR) is a standard objective method for estimating hearing threshold, especially in patients who cannot reliably participate in behavioral audiometry. However, ABR interpretation is usually performed by an expert. This study evaluated whether two general-purpose artificial intelligence (AI) multimodal large language model (LLM) chatbots, ChatGPT and Qwen, can accurately estimate ABR hearing thresholds from ABR waveform images. The accuracy was measured by comparisons with the judgements of 3 expert audiologists.

**Methods:** A total of 500 images each containing several ABR waveforms recorded at different stimulus intensities were analyzed. Three expert audiologists established the reference auditory thresholds based on visual identification of wave V at the lowest stimulus intensity, with the most frequent judgment among the three used as the reference. Each waveform image was independently submitted to ChatGPT (version 5.1) and Qwen (version 3Max) using the same standardized prompt and without additional clinical context. Agreement with the expert thresholds was assessed as mean errors and correlations. Sensitivity and specificity for detecting hearing loss (>20 dB nHL) were also calculated. In cases where the AI and expert thresholds nominally matched, corresponding latency measures were also compared.

**Results:** Auditory thresholds derived from both LLMs correlated strongly with expert opinion, with Pearson *r* = 0.954 for ChatGPT and *r* = 0.958 for Qwen. ChatGPT showed a mean error of +5.5 dB and Qwen showed a mean error of −2.7 dB. Exact nominal agreement with expert values was achieved in 34.6% of ChatGPT estimates and 35.6% of Qwen estimates; agreement within ±10 dB was observed in 75.6% and 80.0% of cases, respectively. For hearing-loss classification, ChatGPT achieved 100% sensitivity but low specificity (20.4%), whereas Qwen showed a more balanced profile with 91.6% sensitivity and 67.5% specificity. Curiously, estimates of wave V latency were markedly poor for both LLMs, with systematic underestimation and weak correlations with the expert judgements.

**Conclusion:** ChatGPT and Qwen demonstrated a moderate ability to estimate ABR thresholds from waveform images, although their performance was not good enough for independent clinical use. Both models captured general patterns of hearing loss severity, but there was systematic bias, limited specificity and sensitivity balance, and poor latency estimation. General-purpose multimodal LLMs may have potential as assistive or preliminary tools, but clinically reliable ABR interpretation will likely require specialized, domain-trained AI systems with expert oversight.

## Introduction

Hearing loss represents a significant global health burden, affecting over 5% of the world’s population and profoundly impacting communication, cognitive function, and quality of life [1,2]. Early and accurate detection of hearing impairment is essential for timely intervention, particularly in populations where behavioral audiometry is impractical, including newborns, young children, and patients who cannot provide reliable behavioral responses. The auditory brainstem response (ABR) has emerged as the gold standard objective measure for hearing threshold estimation in these populations, providing electrophysiological evidence of auditory pathway integrity from the cochlea through to the brainstem nuclei [3,4].

ABR threshold determination relies on the visual identification of wave V (the most robust and clinically relevant component) as stimulus intensities are progressively decreased [5]. However, this identification is inherently subjective and requires considerable expertise. Studies have demonstrated variability in threshold estimation even among experienced audiologists, with inter-rater disagreements particularly common at near-threshold stimulus levels where waveform morphology becomes ambiguous [6,7]. This subjectivity poses challenges for reproducibility across clinical sites and research studies, underscoring the need for more objective and standardized interpretation methods.

Over the past three decades, numerous approaches to automated ABR analysis have been proposed. Traditional statistical methods, including signal-to-noise ratio criteria and cross-correlation techniques, have shown promise but often struggle with low-amplitude responses near threshold [8,9]. More recently, machine learning approaches have demonstrated improved performance. Convolutional neural networks, support vector machines, and ensemble classifiers have been trained to detect the presence or absence of ABR waveforms, with some achieving accuracy comparable to expert human readers [10,11]. Deep learning architectures – including bidirectional long short-term memory networks and vision transformers applied to time-frequency representations – have further advanced the field by capturing complex temporal patterns in ABR signals [12,13]. Despite these advances, most automated systems have been developed and validated on relatively homogeneous datasets, and their generalizability across diverse clinical populations and recording conditions is yet to be evaluated.

The emergence of multimodal large language models (LLMs) capable of processing both text and images has introduced a new paradigm for medical image interpretation [14,15]. Such systems can analyze visual data in conjunction with natural language prompts, potentially offering a flexible and accessible approach to diagnostic tasks. However, evaluations of these models in medical imaging have revealed significant limitations. Studies in radiology, ophthalmology, and pathology have reported highly variable diagnostic accuracy, with performance often substantially below that of specialist physicians and purpose-built convolutional neural networks [16–19]. Notably, these models frequently rely heavily on contextual text information rather than the visual features themselves, raising questions about their true image interpretation capabilities [20].

To date, no study has evaluated the performance of general-purpose multimodal LLMs for ABR threshold estimation. Given the potential for such models to provide rapid, accessible screening tools – particularly in resource-limited settings lacking specialist expertise – an understanding of their current capabilities and limitations is of considerable clinical relevance. Furthermore, as LLM technology continues to rapidly evolve, establishing baseline performance metrics is necessary for tracking progress and identifying areas for improvement. It might even be possible that patients could use LLMs to verify professional diagnoses or clinicians could use them to speed up their work. Could such uses be justified?

The aim of this study was to evaluate and compare the accuracy of two commercially available multimodal LLMs – ChatGPT and Qwen – in estimating hearing thresholds from ABR waveform images, using expert audiologist determinations as the reference standard.

## Methods

This study compared the performance of two large language model (LLM)-based AI chatbots – ChatGPT (OpenAI; multimodal model version 5.1) and Qwen (Alibaba; multimodal model version 3Max) – in estimating hearing thresholds from auditory brainstem response (ABR) waveform images. A dataset of 500 images (each image for a separate ear) of ABR traces for different stimulus intensities was compiled, with expert-determined thresholds serving as the reference standard.

ABR waveforms were extracted from an institutional dataset, without regards to clinical or demographic characteristics. They were all recorded in the same center using an Integrity V500 (Vivosonic Inc., Toronto, Canada) in response to click stimuli presented at descending intensity levels in steps of 10 dB. The stimuli used to evoke ABRs were in the range of 10 to 100 dB nHL. An example is given in Figure 1. Baseline characteristics were not available, as the dataset comprised only ABR waveform images; no clinical or demographic information was used in the analysis by either expert raters or the AI systems.

**Figure 1.**
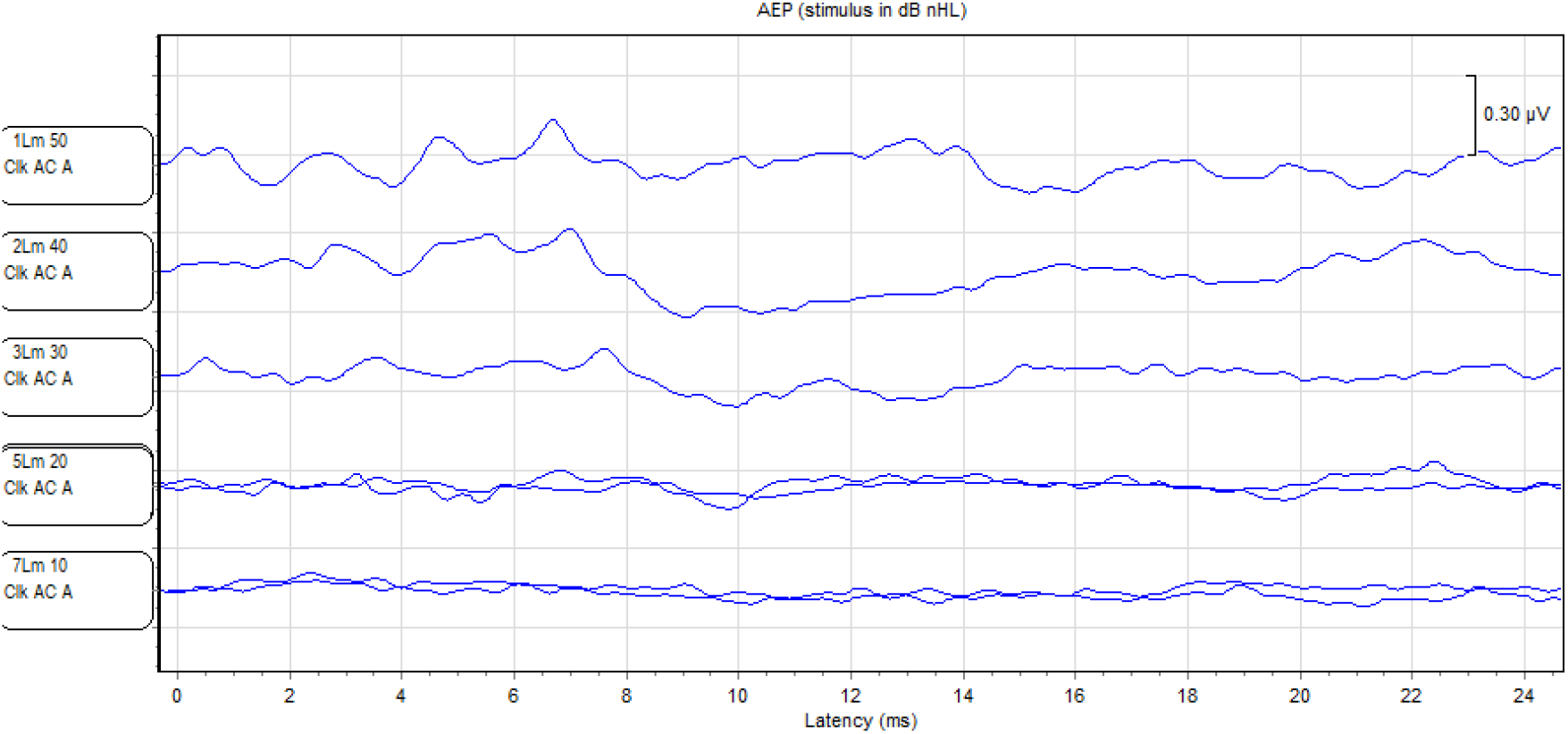
Example of a single ABR image presented to the chatbots, which shows responses from 50 dB down to 0 dB nHL. This is a screen-shot from the system used in our clinic (Integrity V500 from Vivosonic). When ABR waveform V becomes weak and the signal is noisy, standard practice is to record more than one ABR trace at that stimulus intensity; therefore, two traces are shown for 20 and 10 dB nHL.

Initially, sample size was estimated based on reaching an agreement within ±10 dB, treating this endpoint as a single proportion. If we assume an agreement of 80–90% and a 95% confidence interval half-width of ±5%, using the formula n= 1.96^2^×*p*(1-*p*)/*d*^2^ (with *p* = 0.8 to 0.9, and *d* = 0.05), the required then the required sample size was approximately 139–246 images. In this study we used a convenience series of 500 ABR waveform images collected at our center. We expected that the use of 500 samples would improve the precision and robustness of the estimates. Inclusion criteria comprised ABR waveform images for individual ears recorded across multiple stimulus intensities which covered the subject’s hearing threshold; exclusion criteria included recordings with fewer than three intensity levels or otherwise incomplete or technically inadequate data.

Three expert audiologists determined hearing thresholds based on visual identification of wave V at the lowest stimulus intensity level, following standard clinical criteria. Each expert was instructed to provide, for each image, hearing threshold and latency of wave V at threshold. The most frequent result of the three was used as the reference value. All expert evaluations were performed independently to AI-based analysis.

Each ABR waveform image was independently submitted to ChatGPT and Qwen with a standardized prompt requesting a threshold estimate based on the LLM identifying wave V. After each image presentation and response, data was collected in Excel (Microsoft) and the chatbot conversation was reset. Querying of the chatbots took place in November/December 2025. Both systems were provided with the same set of 500 images together with a prompt which provided no additional clinical context. This kept the prompt as simple as possible. After several preliminary tests, we settled on the first simple version which appeared to provide good results. The test indicated that it is crucial to provide the different stimulus levels used to evoke each response.

The prompt used, along with the uploaded image, is set out below for the example shown in Figure 1, which depicts ABR waveforms collected at 50, 40, 30, 20, and 10 dB nHL.

> Analyze the auditory brainstem response (ABR) signals in the attached file, which were recorded at 50, 40, 30, 20 and 10 dB nHL. Identify the lowest intensity level that still shows a clear and replicable ABR waveform (particularly Wave V), and estimate the hearing threshold based on the recordings. Note that there may be multiple traces for each intensity level. Provide only the estimated ABR threshold and the latency of Wave V at that threshold.

Although the prompt requested both threshold and wave V latency, some cases were interpreted by both the raters and the chatbots as showing no identifiable response at any tested intensity. For threshold analysis, such cases were coded as 110 dB nHL. But because wave V latency cannot be assigned to a no-response case, the latency was in these cases treated as missing and excluded from latency analysis.

This research was approved by the Ethics Committee of the Institute of Physiology and Pathology of Hearing (consent no. IFPS:KB/Oświadczenie nr 7/2025 r.).

### Statistical Analysis

Agreement between AI-estimated and expert-determined ABR thresholds was assessed using mean error, mean absolute error (MAE), root mean square error (RMSE), and Pearson’s correlation coefficient. Exact agreement and agreement within ±10 dB and ±20 dB were also calculated. For hearing-loss classification (>20 dB nHL vs ≤20 dB nHL), sensitivity, specificity, and accuracy were calculated for each model, and the models were compared using a McNemar test. Cross-tabulations comparing AI-derived classifications (hearing loss >20 dB nHL vs ≤20 dB nHL) with the expert reference standard were constructed to calculate sensitivity, specificity, and overall diagnostic accuracy for each model. When testing for the accuracy of latency, the latency of wave V was analyzed only in cases where the AI-estimated threshold exactly matched the expert threshold. For these cases, mean error, standard deviation of differences, Pearson’s correlation coefficient, and paired *t*-test results were calculated. A *p*-value < 0.05 was considered statistically significant. No pre-specified subgroup or variability analyses were conducted; any observed variability in model performance (e.g., differences between ChatGPT and Qwen or across threshold ranges) was assessed descriptively and considered exploratory. All analyses were performed in Matlab R2023b (MathWorks, Natick, MA, USA).

## Results

### Hearing threshold detection

Expert evaluations of thresholds based on images of ABR traces at different stimulus intensities were compared with similar evaluations made by the multimodal LLMs. The results are summarized in Table 1. Both LLMs exhibited strong, highly significant correlations with thresholds determined by the experts (Figure 2). With ChatGPT, the Pearson *r* was 0.954 (*p* < 0.001) and with Qwen the figure was 0.958 (*p* < 0.001), indicating that both models were able to accurately capture the overall trend in hearing loss severity.

**Table 1.** Performance comparison of ChatGPT and Qwen for ABR threshold estimation.

| Metric | ChatGPT | Qwen |
| --- | --- | --- |
| N | 500 | 500 |
| Mean Error (dB) | 5.6 | -2.7 |
| MAE (dB) | 8.9 | 8.6 |
| RMSE (dB) | 11.9 | 11.6 |
| Exact numerical match | 34.6% | 35.6% |
| Match within $\pm 10$ dB | 75.6% | 80.0% |
| Match within $\pm 20$ dB | 97.4% | 98.0% |
| Sensitivity (HL detection) | 100.0% | 91.6% |
| Specificity (HL detection) | 20.4% | 67.5% |
| Accuracy (HL detection) | 69.6% | 82.4% |

**Figure 2.**
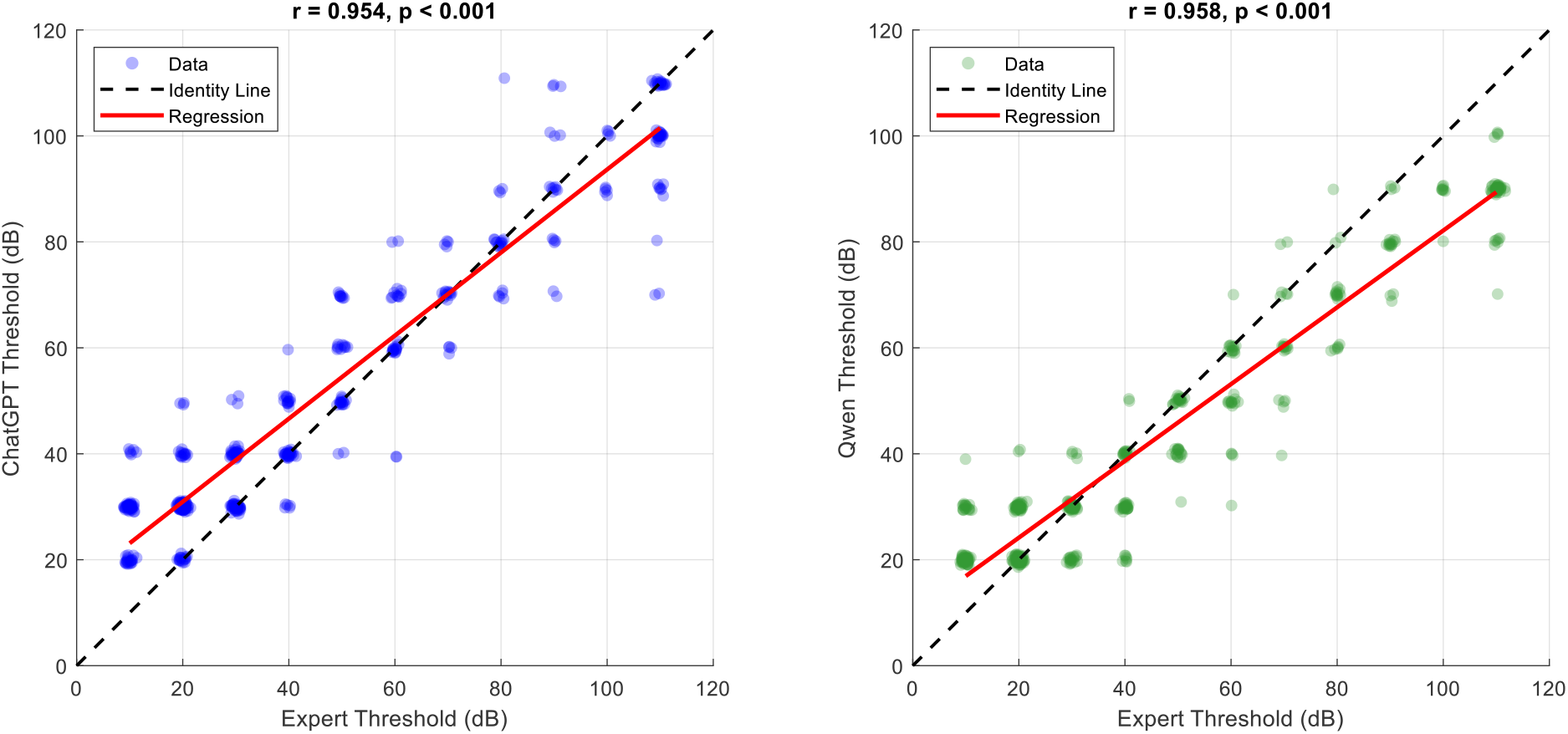
Scatter plots showing correlations between AI model estimates and expert-determined ABR hearing thresholds (click-evoked wave V). Results at left are for ChatGPT and those at right for Qwen. Each point represents one ear (*n* = 500). The data points are slightly offset to visualize multiple points that overlap. The dashed line indicates perfect identity; the solid line represents the fitted linear regression. Pearson correlation coefficient (*r*) and *p*-value are displayed at the top of each panel.

Examination of the trend lines shows that ChatGPT overestimated thresholds relative to the experts by a mean error of +5.5 dB (SD 10.6 dB), with a mean absolute error (MAE) of 8.9 dB. Qwen showed a smaller systematic error of −2.7 dB (SD 11.3 dB) and a slightly lower MAE of 8.6 dB. Exact notional agreement with expert values (effectively ±5 dB) was observed in 34.6% of ChatGPT estimates and in 35.6% of Qwen estimates; estimates within ±10 dB were recorded in 75.6% and 80.0% of cases, respectively.

Next we used the results in Figure 2 to classify whether a person had a hearing loss or not. We looked at all the thresholds which the experts determined were >20 dB nHL, and examined how well the LLMs performed in judging whether that same ear had impaired thresholds. ChatGPT demonstrated perfect sensitivity (100%), but showed markedly low specificity (20.4%), resulting in an overall accuracy of 69.6%. Qwen achieved a more balanced diagnostic profile, with a sensitivity of 91.6% and specificity of 67.5%, yielding superior overall accuracy (82.4%). A McNemar test confirmed that the two LLMs differed significantly in their classification decisions (*χ*^2^(1) = 32.01, *p* < 0.001), with Qwen demonstrating more clinically balanced performance across both diagnostic categories (hearing loss or not).

### Detection of wave V latency at threshold

Wave V latencies at hearing threshold as determined by ChatGPT and Qwen were then compared against the expert’s assessments. Analysis was restricted to cases where the AI-determined hearing threshold matched the expert threshold exactly (since it is known that latency depends strongly on hearing thresholds). Moreover, as indicated earlier, in those few cases where no wave V was detected (which happened only for ChatGPT), these cases were also excluded.

For matched-threshold pairs, ChatGPT showed a mean wave V latency error of −0.87 ms (SD = 1.14 ms) and Qwen −1.49 ms (SD = 1.12 ms) relative to expert readings (Table 2). Both these results were significantly different from those of the experts (bottom row of Table 2). Pearson correlations for latencies (Figure 3) were much weaker than for thresholds, with *r* = 0.26 (*p* = 0.001) for ChatGPT and *r* = 0.20 (*p* = 0.0076) for Qwen. Thus, both LLMs systematically underestimated wave V latency compared to the experts.

**Table 2.** Performance comparison of ChatGPT and Qwen for ABR latency estimation. Result of *t*-test comparison with expert determined latency provided in the last line. For ChatGPT, the number of valid latency pairs was smaller than the number of exact threshold matches because some exact matches corresponded to no-response cases, for which latency was unavailable.

| Metric | ChatGPT | Qwen |
| --- | --- | --- |
| Threshold matches ($n$) | 173 | 178 |
| Threshold match rate (%) | 34.6 | 35.6 |
| Valid latency pairs ($n$) | 153 | 178 |
| Mean error (ms) | -0.868 | -1.487 |
| SD of differences (ms) | 1.136 | 1.120 |
| $t$-statistic | -9.443 ($p < 0.001$) | -17.725 ($p < 0.001$) |

**Figure 3.**
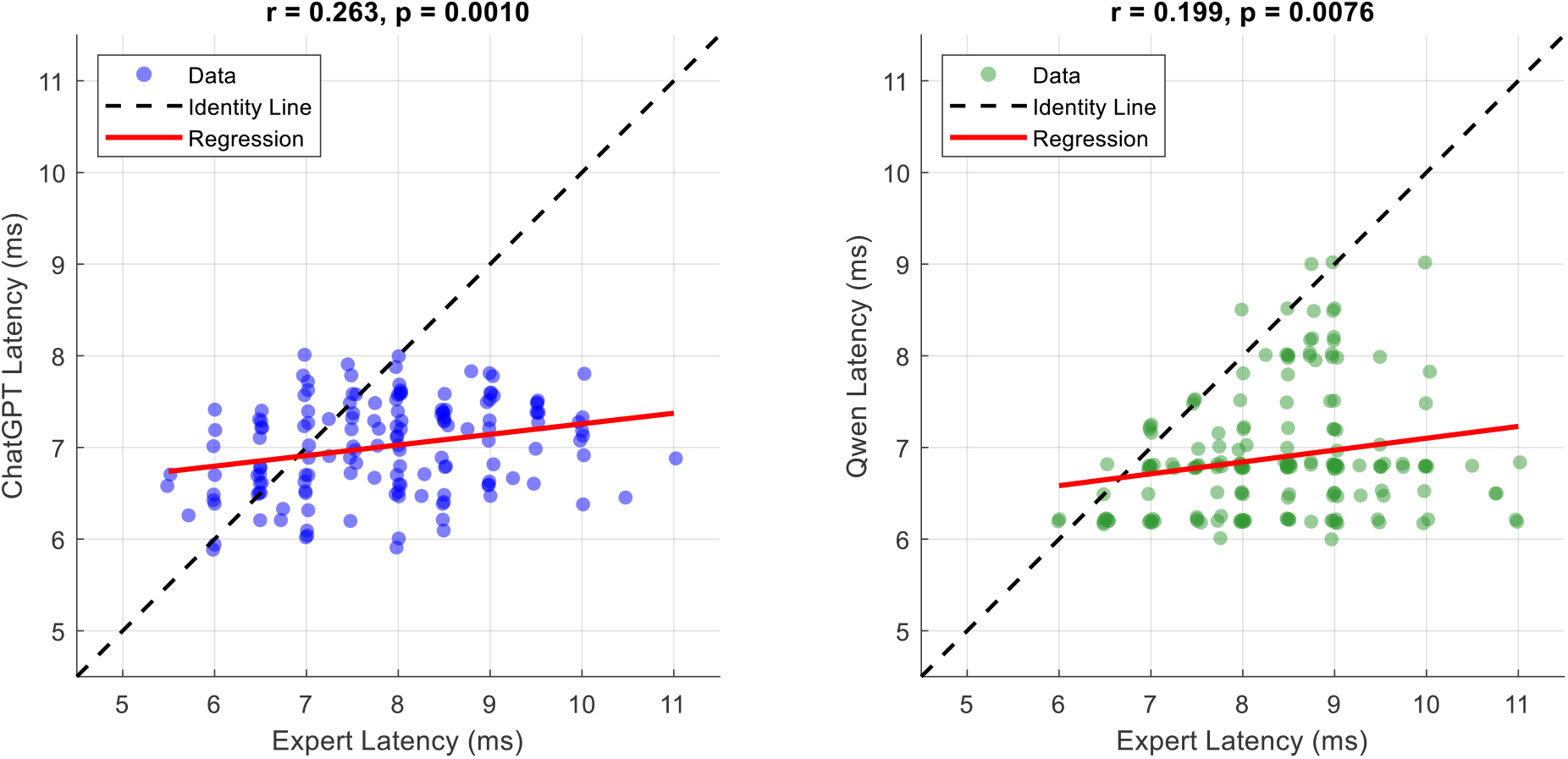
Scatter plots of wave V latencies determined by ChatGPT (left) and Qwen (right) against expert reference latencies, restricted to cases where the AI-determined hearing threshold matched the expert threshold. The data points are slightly offset to visualize multiple points that overlap. The dashed line indicates perfect identity; the solid line represents the fitted linear regression. Pearson correlation coefficient (*r*) and *p*-value are displayed at the top of each panel.

## Discussion

To our knowledge, this study is the first systematic evaluation of general-purpose multimodal LLMs, specifically ChatGPT and Qwen, for hearing threshold estimation based on ABRs. Both models showed strong correlations with expert-derived ABR thresholds, indicating that they were able to capture certain clinically relevant aspects of waveform morphology. However, neither model achieved error levels low enough for clinically reliable use. ChatGPT showed a consistent tendency to overestimate thresholds, whereas Qwen produced estimates that were slightly better but on the low side. However, wave V latencies were substantially poorer for both models, with low correlation coefficients, large discrepancies relative to expert assessments, and considerable variability.

These findings should first be interpreted in the context of human inter-rater variability. Surprisingly, despite the central role of ABR in infant audiological diagnosis, the literature on agreement between expert ABR raters is limited. The field would benefit from large, multicenter studies using standardized clinical protocols in human patients. The available evidence is mixed. Vidler and Parker [6] reported discrepancies between experienced clinicians of up to 60 dB. On the other hand, some studies have shown relatively good agreement, including a correlation of 0.873 reported by Zaitoun et al. 2016 [21] and 93.2% agreement within a 10 dB limit reported by Gos et al. 2025 [22]. The values from the latter study are better than the agreement observed here for the LLMs, which reached 76% and 80% within ±10 dB.

The present results also compare unfavorably with AI systems developed specifically for ABR analysis. Previous studies using convolutional neural networks, ensemble classifiers, and other purpose-built approaches have reported agreement rates exceeding 85–90% within ±10 dB of expert thresholds [10,11]. More recently, ‘ABRpresto’ achieved 92% agreement within ±10 dB [23], while an older system based on statistical methods and signal processing achieved 94% agreement [24]. The mean absolute errors observed in the present study – 8.9 dB for ChatGPT and 8.6 dB for Qwen – are clinically important because errors of this magnitude may lead to misclassification of hearing status and inappropriate management decisions. Nevertheless, the relatively strong correlations suggest that both models acquired a limited ability to interpret ABR waveform morphology, which is notable given that neither was designed or trained specifically for this task.

The results that most strongly undermine the usefulness of these LLMs are those which involve latency estimation. The large discrepancy in latency relative to experts, often about 1 ms or more (Figure 3), suggests that the chatbots may have identified a waveform component other than wave V. Some previous work on specialized AI systems has reported agreement with experts within approximately ±0.1 ms for latency estimation [25]. This marked difference is important because it suggests that even when threshold estimates appear reasonably close to those of experts, they may be based on incorrect physiological interpretations of the waveform. In other words, we are suggesting that apparent agreement in threshold estimates may possibly arise from detecting the wrong waveform feature.

The performance of the two general-purpose multimodal models we tested is also consistent with findings from other medical imaging domains. Studies evaluating multimodal ChatGPT in radiology, ophthalmology, and dermatology have generally found performance below that of specialists and below that of dedicated AI systems [17–19]. Our findings extend these observations to electrophysiological waveform interpretation and suggest that the current generation of multimodal LLMs lack the precision necessary for quantitative medical measurements.

An interesting finding of this study was the different error patterns observed for the two models. ChatGPT showed a systematic tendency toward overestimation, with a mean error of +5.5 dB, whereas Qwen showed the opposite, with a mean error of −2.7 dB. These opposing biases have distinct clinical implications.

ChatGPT’s overestimation tendency resulted in very high sensitivity (100%) but poor specificity (20.4%) for the detection of hearing loss. In a screening context, this behavior would lead to substantial over-referral of normal-hearing individuals for further evaluation. Although inefficient and resource-intensive, such an error pattern is generally less harmful than missed pathology. In contrast, Qwen’s underestimation pattern yielded better specificity (67.5%) but lower sensitivity (91.6%). Clinically, this is more concerning, as missed hearing loss may delay intervention during critical periods of auditory and language development, particularly in children.

These contrasting behaviors may reflect differences in training data, model architecture, or internal calibration. ChatGPT’s tendency to err toward pathology when uncertain may represent a more conservative response profile, whereas Qwen’s greater tendency to normalize findings may reflect a different optimization strategy or training distribution. At present, however, such explanations remain speculative because the internal decision processes of these commercial systems are not accessible.

Several factors may explain the limitations observed in this study. First, ABR waveform interpretation requires the identification of subtle morphological features, particularly the presence and replicability of wave V, which may be difficult for models trained predominantly on natural images and photographs. Near-threshold ABRs are especially challenging because wave V emerges gradually from background noise, making the task fundamentally different from conventional image-recognition problems.

Second, multimodal LLMs have been shown to rely heavily on textual context when both image and text are available [20]. In the present study, only minimal prompts were provided, which may have forced greater reliance on image analysis. However, without access to model internals, it is impossible to determine to what extent these systems truly analyzed the waveform morphology as opposed to applying general heuristics.

Third, the training data of these general-purpose models likely included very few, if any, ABR images. Unlike radiographs or retinal images, ABR waveforms belong to a highly specialized domain with relatively little publicly available labeled data. This scarcity of domain-specific training material likely constrains performance, regardless of the general capabilities of the underlying model.

Despite these limitations, the evaluated LLMs demonstrated a meaningful capacity for ABR interpretation. This is noteworthy, as ABR threshold assessment is not yet widely automated and still largely depends on expert judgment, especially near threshold. It is therefore striking that general-purpose multimodal systems, without explicit domain-specific training, achieved agreement with expert assessments within ±10 dB in approximately 76–80% of cases. Moreover, the input data were not specially formatted or optimized for AI analysis. Instead, as illustrated in Figure 1, the models were given relatively raw screenshots captured directly from the recording system. In many recordings, repeated measurements at the same stimulus intensity produced overlapping waveforms, adding further complexity to interpretation. Even under these conditions, both ChatGPT and Qwen were often able to produce threshold estimates reasonably close to those of expert audiologists. This observation raises the possibility that commercial multimodal models may be capable of handling ABR recordings generated by other systems as well, although this remains unproven because no external validation was performed. Nevertheless, the present findings also make clear that these models are not yet suitable for professional application. Their outputs were clearly non-random, but they remain insufficiently accurate and reliable for clinical or research contexts in which precise interpretation is essential.

In principle, such models might have limited utility in resource-constrained settings that lack specialist expertise, provided that their outputs are treated strictly as preliminary and always verified by qualified clinicians. ChatGPT’s very high sensitivity suggests possible value as a conservative screening aid. However, even in that context, the current error rate, and occasional very large discrepancies, remains problematic. Similarly, for research applications involving large-scale ABR analysis, these systems might assist with preliminary annotation or triage, but only when combined with expert review, especially for borderline or uncertain cases.

Taken together, the results support two main conclusions. First, it is impressive that general-purpose multimodal LLMs can produce plausible evaluations in a specialized domain such as ABR threshold detection, despite high inter-patient variability, differences between acquisition systems, and the lack of universal standards for ABR presentation. Second, this performance remains clearly inferior to that of specialized systems and far below the level needed for dependable professional use. Therefore, although commercial multimodal LLMs may be interesting as exploratory tools or for preliminary assessment, future progress in this area should focus on the development of smaller, specialized AI systems trained specifically for ABR analysis. Such systems are more likely to achieve the precision required in practice and may also offer an important advantage in allowing local deployment, thereby reducing concerns related to the transfer of sensitive medical data.

The study has several limitations relevant to STARD-AI reporting. It was conducted at a single center using screenshots from one ABR acquisition system rather than raw electrophysiological signals, and no external validation dataset was used. The sampling strategy and case mix may therefore limit generalizability to other populations, recording protocols, and display formats.

## Conclusions

ChatGPT and Qwen demonstrate moderate but imperfect accuracy for ABR threshold estimation from waveform images, with systematic errors that differ between systems. Current performance falls short of clinical requirements, with approximately 20–24% of estimates exceeding the ±10 dB agreement margin. While these general-purpose multimodal LLMs show promise as assistive tools, substantial improvements in accuracy and reliability are needed before consideration for clinical implementation. Domain-specific training and careful integration with expert oversight represent promising strategies for future development.

## Data Availability

All data produced in the present study are available upon reasonable request to the authors.

## Acknowledgement

The authors thank Elżbieta Gos, Małgorzata Pastucha, Marietta Reck, and Natalia Usarek for help with data preparation and collection, and Andrew Bell for comments on an earlier version of this manuscript. ChatGPT (OpenAI) was used for initial brainstorming and language suggestions; all text was subsequently revised by the authors and a native English-speaking editor to ensure accuracy and clarity. This study was carried out within the framework of the grant “ABR.AI – An AI-based system for estimating hearing thresholds from intensity series of wave V responses” (no. 2024/ABM/03/KPO/KPOD.07.07-IW.07-0222/24-00), funded by the Medical Research Agency (Agencja Badań Medycznych, ABM).

## Notes

### Competing Interest Statement

The authors have declared no competing interest.

